# Mendelian randomization confirms the role of Y-chromosome loss in Alzheimer’s Disease etiopathogenesis in males

**DOI:** 10.1101/2022.07.20.22277657

**Authors:** Pablo García-González, Itziar de Rojas, Sonia Moreno-Grau, Laura Montrreal, Raquel Puerta, Emilio Alarcón-Martín, Inés Quintela, Adela Orellana, Victor Andrade, Pamela Martino Adami, Stefanie Heilmann-Heimbach, Pilar Gomez-Garre, María Teresa Periñán, Ignacio Alvarez, Monica Diez-Fairen, Raul Nuñez Llaves, Claudia Olivé Roig, Guillermo Garcia-Ribas, Manuel Menéndez-González, Carmen Martínez, Miquel Aguilar, Mariateresa Buongiorno, Emilio Franco-Macías, Maria Eugenia Saez, Amanda Cano, Maria Bullido, Luis Real, Eloy Rodríguez-Rodríguez, Jose Royo, Victoria Álvarez, Pau Pastor, Gerard Piñol-Ripoll, Pablo Mir, Miguel Calero Lara, Miguel Medina Padilla, Pascual Sánchez-Juan, Angel Carracedo, Sergi Valero, Isabel Hernandez, Lluis Tàrraga, Alfredo Ramirez, Mercé Boada, Agustín Ruiz

**Author notes:** **<u>CORRESPONDING AUTHOR</u>**: Pablo García González, **ADRESS**: Marques de Sentmenat 57, Barcelona, Spain 08029.

## Abstract

Mosaic loss of chromosome Y (mLOY) is a common ageing-related somatic event occurring exclusively in men and has been previously associated with Alzheimer’s disease (AD). However, mLOY estimation from genotype microarray data only reflects the mLOY degree of subjects at the moment of DNA sampling. Therefore, mLOY phenotype associations with AD can be severely age-confounded in the context of genome-wide association studies. Here, we applied Mendelian randomization to construct an age-independent polygenic risk score of mLOY (mloy-PRS) using 114 autosomal variants. The mloy-PRS instrument was associated with an 80% increase in mLOY risk per SD unit (p=4.22·10^−20^) and was orthogonal with age. We found that a higher genetic risk for mLOY was associated with faster progression to AD in males with mild cognitive impairment (HR=1.23; p=0.01). Importantly, mloy-PRS had no effect on AD conversion or risk in the female group. The male-specificity of the observed effects suggests that these associations of mLOY with AD are caused by the inherent loss of the Y chromosome, and not by the increased genomic instability underlying mLOY risk. Additionally, we found that blood mLOY phenotype was associated with increased CSF levels of total tau and phosphorylated tau181 in subjects with mild cognitive impairment and dementia. Our results strongly suggest that mLOY is involved in AD pathogenesis. Furthermore, we encourage researchers to use this mloy-PRS instrument to find unbiased associations between mLOY and ageing-related diseases.

## INTRODUCTION

Alzheimer’s disease (AD) is the leading cause of dementia worldwide, accounting for 60–80% of total cases [1]. While Mendelian inheritance is suspected to cause early onset AD (<65 years) [2], late onset AD (LOAD, >65 years) is a complex, multifactorial disease influenced by both genetic factors and life exposures. The genetic contribution to LOAD is estimated to be 60–80% [3], being *APOE* the most prominent locus discovered to date [4]. However, demographic features also play a predominant role in AD. Notably, age is considered the most important risk factor for LOAD [5]. On the other hand, women represent nearly two thirds of the global population with AD [1] and show higher rates of cognitive decline [6, 7] than men. However, whether sex should be considered a risk factor for AD or instead a source of disease heterogeneity is a matter of intense debate [8]. Recent reviews highlighted the importance of reporting results for sex interactions and sex-stratified AD data instead of the more widely used approach of adjusting data by sex [9]. These approaches may help elucidate differences in sex-specific AD risk profiles, which will be of great value in the incoming age of precision medicine.

The male-specific region of chromosome Y is one of the most unexplored regions of the human genome and has long been considered a genetic wasteland. Mosaic loss of chromosome Y (mLOY) in blood cells is the most common known form of somatic mosaicism in humans [10–12]. Genetic factors together with age, smoking and other environmental stressors are well known risk factors for mLOY [13]. Genetic variants associated with mLOY risk are mainly related to mitotic processes, cell cycle regulation, DNA damage sensing and response, and apoptotic processes [14]. mLOY was initially considered a phenotypically innocuous, age-related trait [15–18]. However, there is increasing evidence that mLOY in blood cells has a direct effect in the etiopathogenesis of several diseases affecting different tissues. Specifically, blood cell mLOY has been associated with susceptibility to multiple ageing-related diseases, i.e., AD [19], non-hematological cancer [10, 20], cardiovascular diseases [21, 22] and all-cause mortality risk [10]. The main proposed mechanism to explain blood mLOY pathogenesis is by impairment of immune functions caused by the loss of the Y chromosome in leukocytes [23–25]. However, it has been described that autosomal genetic predisposition for mLOY is associated with breast cancer in women, indicating that the underlying genomic instability can also explain the associations between mLOY and disease risk [14].

Here, we aimed to study mLOY’s impact on AD risk in the GR@ACE and Dementia Genetics Spanish Consortium (DEGESCO) cohorts [26, 27]. First, we checked for blood mLOY associations with AD in both a case-control setting and in the phenoconversion process from mild cognitive impairment (MCI) to all-cause dementia and AD. Subsequently, to remove age confounding effects, we generated an autosomal, age-independent polygenic risk score (PRS) of mLOY and analysed its effect on AD status and progression in both sexes. Finally, we analysed mLOY’s impact in different AD-related biomarkers in CSF.

## METHODS

### The GR@ACE-DEGESCO cohort

The GR@ACE-DEGESCO cohort comprises AD patients and controls from the Spanish population. AD patients were collected from ACE Alzheimer Center Barcelona and 12 other cohorts included in the Dementia Genetics Spanish Consortium (DEGESCO) (Supplementary Table 1). Control individuals were provided by ACE Alzheimer Center (Barcelona, Spain), Valme University Hospital, the Spanish National DNA Bank Carlos III (Salamanca, Spain) and other DEGESCO members. DNA extracted from peripheral blood or saliva (Supplementary Table 1) was genotyped in the Spanish National Center for Genotyping (CeGen, Santiago de Compostela, Spain), using the Axiom 815K Spanish Biobank Array (Thermo Fisher), as previously described [26, 27].

### The ACE MCI-EADB cohort

The EADB cohort is a prospective cohort conformed by MCI individuals recruited between 2006 and 2013 at ACE Alzheimer Center Barcelona. Briefly, individuals with a Clinical Dementia Rating Scale (CDR) of 0.5 and older than 60 years were selected and underwent at least one follow-up, consisting of neurological, neuropsychological and social work evaluations. Detailed definition of the ascertainment of this cohort has been described [28, 29]. DNA genotyping was performed as described elsewhere [30]. Briefly, DNA extracted from peripheral blood was genotyped with the Illumina Infinium Global Screening Array (GSA, GSAsharedCUSTOM_24+v1.0) in LIFE & BRAIN CENTER, (EADB node, Bonn, Germany) and SNP genotype calls were obtained from raw probe intensity data in the same center.

### Criteria for AD diagnosis, case-control setup

AD diagnoses were established in all cases by a multidisciplinary working group conformed by neurologists, neuropsychiatrists and social workers, following DSM-IV criteria for dementia and the National Institute on Aging and Alzheimer’s Association’s (NIA-AA) 2011 guidelines for AD definition. In the present study, individuals were labeled as AD when possible or probable AD was endorsed by neurologists at any point of their clinical history. Written informed consent was obtained from all participants. The Ethics and Scientific Committees have approved this research protocol (Acta 25/2016, Ethics Committee. H., Clinic I Provincial, Barcelona, Spain).

### Assessment of MCI to dementia/AD conversion

MCI to dementia conversion was determined by integrating CDR, Global Deterioration Scale (GDS), and diagnostic assessments at ACE Alzheimer Center Barcelona, assigned at a consensus conference including neurologists, neuropsychologists and social workers [31]. Conversion to dementia was defined as the first clinical evaluation reporting a diagnosis of AD [32, 33], vascular dementia [34], mixed dementia (AD with cerebrovascular disease), frontotemporal dementia [35, 36] or dementia with Lewy bodies [37], combined with a CDR change from 0.5 to >=1, and GDS >=4. AD converters were defined as the fraction of converters to dementia that were diagnosed with AD. Baseline criteria varied depending on whether the exposure was mLOY phenotype or it’s associated PRS: In the first case, baseline was defined as the moment of blood sampling used afterwards for germline DNA extraction, genome-wide genotyping and mLOY estimation. We selected only those individuals who met Petersen’s criteria [38, 39], for amnestic and non-amnestic MCI, at the closest clinical evaluation to DNA sampling. Because genotypes used for PRS estimates are invariable, baseline was defined as the patient’s first clinical record meeting Petersen’s criteria for PRS analysis. Follow-up time was defined as the time window between baseline and: (a) the date of conversion to dementia (converters) and (b) the date of last clinical evaluation (non-converters). In order to have a prospective cohort, disease progression models only included individuals who were either originally selected as controls/MCIs in the GR@ACE-DEGESCO case-control cohort, or present in the MCI cohort (ACE MCI-EADB).

### LOY determination

PennCNV [40–42] was used to process CEL files, following the recommended workflow for Affymetrix arrays [43] to obtain Log R Ratio (LRR) and B allele frequency (BAF) values for each array probe in our dataset. Determination of mLOY was performed using MADloy package for R [44]. Briefly, this method estimates mLOY by normalizing the median LRR of probes found at the male specific region of chromosome Y (mLRR-Y), against the 5% trimmed mean LRR of autosomal chromosomes. Thus, only probes located between pseudoautosomal regions 1 (PAR1) and 2 (PAR2) in chromosome Y, excluding the X transposed region, (chrY:6611498-24510581; hg19/GRCh37) are used to compute mLRR-Y. To call mLOY status, we used the mLRR-Y_thres_ method of the MADloy package. Briefly, a threshold is determined by extrapolating the 99% confidence interval of the positive side of the cohort mLRR-Y distribution [10]. Then, samples with mLRR-Y values below the empirically calculated threshold are assigned mLOY status (or calls). To overcome computational power limitations, mLOY calls were obtained in two randomized batches. B-deviation (Bdev), defined as the mean deviation from the expected BAF (0.5) for heterozygous SNPs, located in PAR1 regions (PAR1-Bdev) was used as a complementary indicator of mLOY (Supplementary Figure 1).

### Sample processing and QC

LRR and BAF values for all biallelic markers were obtained from 20 068 CEL files (call rate >0.97 per sample and >0,985 per plate). Then, reported male samples were retrieved, and samples with mean LRR-X and LRR-Y corresponding to female (XX) or sexual chromosome aneuploidies were discarded. Additionally, samples with high heterozygosity rate, high chromosome X heterozygosity and population outliers were removed from our dataset. Samples with LRR SD > 0.46, a standard QC parameter for Affymetrix LRR data, were removed. GENESIS R package [46] was used to examine relatedness within our dataset. Second degree relatives were detected by using a kinship threshold of 0.046875 and were filtered out of the dataset. Finally, outliers in the mLRR-y and b-deviation distribution were removed. Specific QC procedures and sample filtering steps for each analysis are summarized in Supplementary Figure 2.

### mLOY autosomal Polygenic Risk Score

Processing, QC and imputation of the genome-wide SNP data were performed as described elsewhere [27, 30]. An mLOY autosomal polygenic risk score (mloy-PRS) was calculated based on independent genome-wide significant variants previously described [14]. Out of the 156 reported SNPs, we excluded those unavailable in our dataset, considered rare variants (MAF<0.01), with low imputation quality (R^2^<0.3), or located within the sexual chromosomes, leaving us with a final number of 114 autosomal SNPs (Supplementary Table 2). mloy-PRS was calculated for all individuals in the GR@ACE-DEGESCO and ACE MCI-EADB cohorts by adding the dosage of risk alleles weighted by their reported effect sizes (beta coefficients). To ease interpretation of results, mloy-PRS units were standardized (SD=1).

### Core AD biomarkers & targeted proteomics

Levels of Abeta-42, tau phosphorylated at position 181 (p-tau) and total tau were measured the same day in cerebrospinal fluid (CSF) samples obtained via lumbar punctures using commercially available ELISA immunoassays (INNOTEST® β-AMYLOID (1-42), INNOTEST® hTAU, and INNOTEST® PHOSPHO-TAU(181P) (Fujirebio, Spain).

CSF and paired plasma samples collected the same day, as described elsewhere [47, 48], underwent targeted proteomics using ProSeek® multiplex immunoassay by Olink Proteomics (Uppsala, Sweden). Protein concentration was measured for 184 proteins included in the commercially available ProSeek® Multiplex panels (Inflammation & Neurology) in both fluids. Quality control details and further description of this data are provided elsewhere [49].

### Statistical Analysis

Data processing and analysis was performed with R Software [50]. To harmonize effect directions, the mLRR-Y variable was multiplied by -1 due to lower values of mLRR-Y representing a higher degree of mLOY. Logistic regressions adjusted by age, *APOE* genotype and population structure were fitted for case-control analysis. Survival R package [51] was used to fit Cox proportional-hazards models to assess MCI conversion to all-cause dementia or AD. Due to the age-dependent nature of mLOY [11, 52], only individuals with available age at DNA sampling information were used for analyses involving mLOY or mLRR-Y. To correct for population structure, only the principal components (PCs) that were associated with the dependent variable were included in the models. Population structure was not corrected in the Cox models as PCs showed no effect on disease progression (Supplementary Table 3, Supplementary Figure 3), likely because all MCI samples came from the same center (ACE Alzheimer Center Barcelona). *APOE* genotypes were modelled as a continuous variable ranging from -2 to 2, where each *APOE*-ε2 allele contributed with -1 and each *APOE*-ε4 allele added +1, as previously described [29]. In order to control ascertainment and genotyping bias between MCI cohorts (GR@ACE-DEGESCO & ACE MCI-EADB), a dichotomic variable was introduced in models testing association of mloy-PRS with disease progression. For analysis of Olink proteomic data, linear regressions were adjusted by age, the time window between DNA sampling and lumbar puncture, and *APOE* genotype. Due to the high correlation between many CSF proteins, total tau and p-tau levels (Supplementary Figure 4), we also included models adjusted by total tau levels. Fixed-effect inverse variance weighted meta-analysis was performed with the rma.uni function included in the metafor R package [53].

## RESULTS

We computed mean LRR-Y and LRR-X values for 7 954 clinically reported male samples and plotted these values, generating clusters that allowed us to check the sexual chromosome dosages of these individuals (Supplementary Figure 5). We detected one individual with a gain of chromosome Y (GOY, XYY) compatible with a supermale syndrome and three individuals with Klineffelter syndrome (XXY). Women (XX), GOY, Klineffelter individuals, and outliers were discarded prior to mLOY computation. For the 7 843 remaining XY individuals, we removed second-degree or lower relatives, and samples with low genotype call rate (≤0.97) or excess heterocigosity (>3SD over cohort mean heterocigosity). We ran principal component analysis to identify the population structure and removed 72 individuals from non-European population (>6SD from 1000 Genomes European population mean). Subjects with detectable autosomal chromosomopathies, i.e., Down’s Syndrome, were also excluded. After applying these exclusion criteria, we split the remaining 6 955 male samples in two randomized batches and performed mLRR-Y computation and mLOY calling. We did not detect batch effect due to cohort splitting (Supplementary Figure 6). We excluded 12 additional samples with LRR SD > 0.46. Finally, we plotted mLRR-Y and PAR1-Bdev values in order to identify and remove individuals with detectable anomalies in chromosome Y (i.e. partial loss of chromosome Y) or loss of heterocigosity in the PAR1 region (Supplementary Figure 5). QC and filtering steps for analysis of mLOY phenotypes and mloy-PRS of our data are summarized in Supplementary Figure 2.

To have a first glimpse at our data, mLRR-Y values of all AD cases and controls were plotted with respect to age (Figure 1). The first thing that became apparent was that our control population is significantly younger than the AD population. Additionally, mLOY occurrence before 65 years was a very rare event in our cohort, indicating that our control population below this age threshold may not be representative for assessing mLOY’s effect on AD. Moreover, our control population mostly lacked individuals older than 85 years. Consequently, we decided to stablish a 65–85 age window for analyzing mLOY’s effect on AD. This matches the usual age at onset range for preclinical, prodromal, and mild dementia stages for LOAD in our population [31] and helped reduce the age gap between our case and control groups (Supplementary Table 4). Concordant with previous reports, we observed a clear age-related increase of mLOY events in the older individuals (Figure 1). Age was associated with mLOY occurrence in males aged 65–85, with an estimated 1% increase in the chance of developing LOY every year (p=3.50·10^−11^).

**Figure 1.**
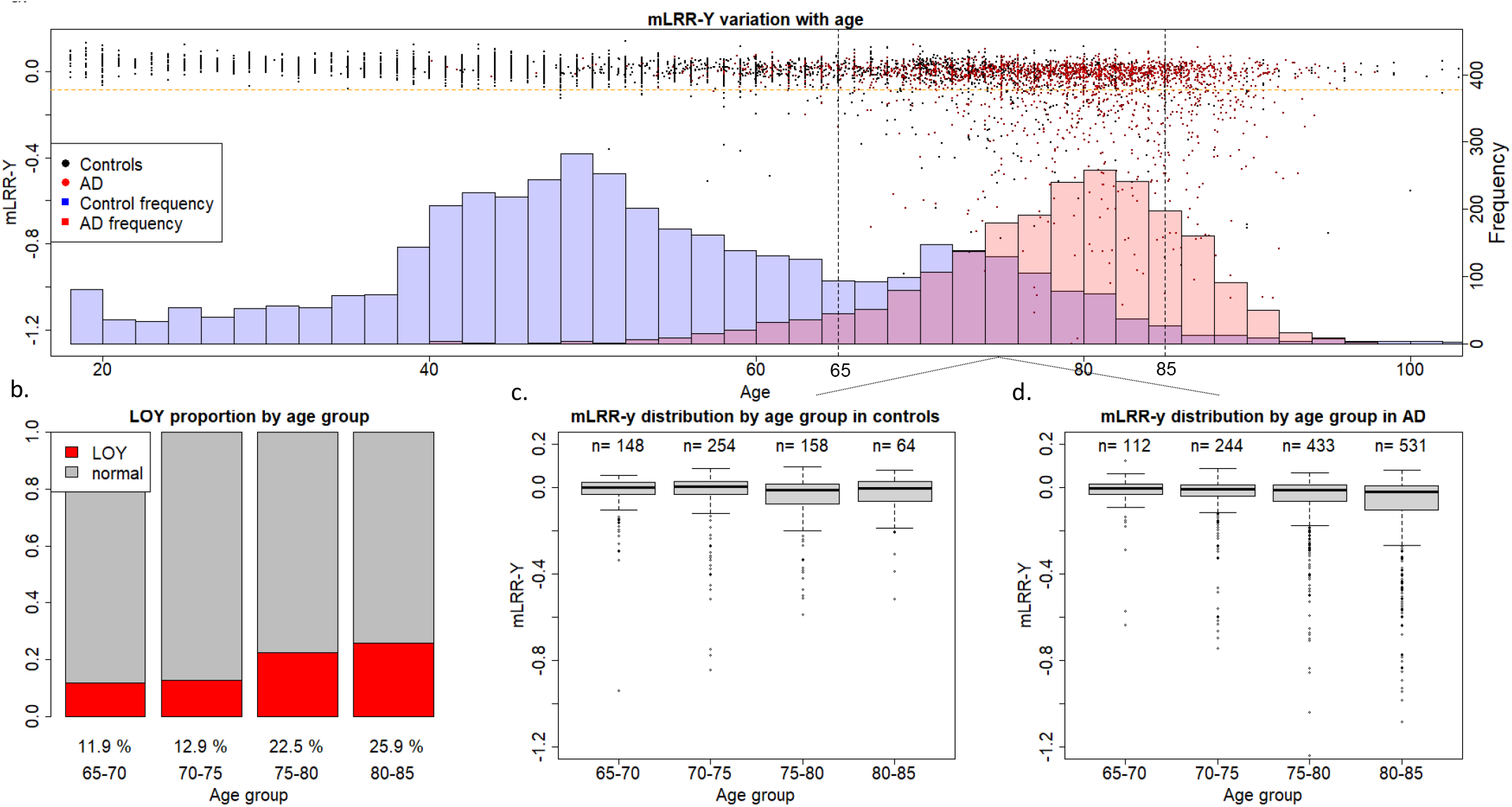
mLRR-y variation with age in the GR@ACE-DEGESCO cohort. **a**. Age and mLRR-Y distribution in the case and control groups. The dots represent mLRR-Y values for individual samples and the histogram represents the age distribution across the case and control groups **b**. Proportion of individuals with mLOY in the different age groups based on age at blood sampling. **c–d** mLRR-Y distriab.ution for the different age groups based on age at blood sampling in control and AD individuals, respectively.

Then, we assessed if continuous mLRR-Y values were differentially distributed amongst cases and controls. Both unadjusted Kolmogorov-Smirnoff test (D=0.18124; p < 2.2·10^−16^) and ANCOVA adjusted by age at DNA sampling and *APOE* (F=68.0; p=2.54·10^−16^, Supplementary Table 5) yielded highly significant results in models including all available males in our cohort.

Next, we fitted logistic regressions for AD status using mLOY calls and mLRR-Y, defining 3 experimental setups: a) all men with available age at DNA sampling, b) 65– 85-year-old men and c) dividing the data in age groups (65–70, 70–75, 75–80 & 80–85 years old). Logistic regressions were adjusted by age at blood sampling, *APOE* genotype and relevant PCs (Supplementary Table 6).

We found that the continuous mLRR-Y variable was associated with AD in the group including all men (N=2 697; OR=2.74; p=0.01), indicating that AD cases had an increased degree of LOY mosaicism compared to controls. We also observed increased mLOY levels in AD males in the 65–85 (N=1 944; OR=2.19; p=0.09) and stratified age groups (Table 1) with respect to controls, but these differences were not statistically significant. Importantly, we noticed that the significant effect observed in the model including all available males could be, at least partially, driven by the dramatic age differences between AD cases and controls in our cohort (Figure 1, Supplementary Table 4), even after adjusting our data by age. mLOY calls were not significantly associated to AD in the group including all men (N=2 697; OR=1.14; p=0.35), the 65– 85-year-old group (N=1 944; OR=1.04; p=0.81), nor in the age-stratified groups (Table 1).

**Table 1.** Logistic regression results using mLRR-Y and mLOY calls as predictors for AD status. 3 experimental setups were defined: a) all men with available age at DNA sampling, b) 65–85-year-old men, and c) a stratification of 65–85-year-old men in age groups (65–70, 70–75, 75–80, & 80–85 years old). Models were adjusted by age at blood sampling, *APOE* genotype, and PCs. In the age-stratified models, only the effect of mLRR-Y or mLOY are displayed.

|  |  | All available males |  |  |  |  | 65-85 year-old males |  |  |  |  | Stratified 65-85 year-old males |  |  |  |  |  |
| --- | --- | --- | --- | --- | --- | --- | --- | --- | --- | --- | --- | --- | --- | --- | --- | --- | --- |
|  |  | OR | SE | P | CI2.5 | CI97.5 | OR | SE | P | CI2.5 | CI97.5 | Age group | OR | SE | P | CI2.5 | CI97.5 |
| mLRR-Y | mLRR-Y | 2.74 | 0.4 | 1.13·10<sup>-2</sup> | 1.29 | 6.16 | 2.19 | 0.47 | 9.28·10<sup>-2</sup> | 0.18 | 1.1 | 65-70 | 1.47 | 1.43 | 0.79 | 0.07 | 21.96 |
|  | Age | 1.16 | 0.01 | 2.74·10<sup>-104</sup> | 1.15 | 1.18 | 1.27 | 0.01 | 6.61·10<sup>-70</sup> | 1.24 | 1.31 | 70-75 | 1.74 | 0.77 | 0.48 | 0.39 | 8.18 |
|  | APOE | 2.52 | 0.08 | 1.61·10<sup>-32</sup> | 2.17 | 2.94 | 2.86 | 0.10 | 5.40·10<sup>-28</sup> | 2.38 | 3.46 | 75-80 | 2.37 | 0.77 | 0.27 | 0.57 | 12.16 |
|  | PC1 | 2.01·10<sup>3</sup> | 2.6 | 3.39·10<sup>-3</sup> | 12.48 | 3.29·10<sup>5</sup> | 0.01 | 3.33 | 0.11 | 0.00 | 3.48 | 80-85 | 44.44 | 2.51 | 0.13 | 0.99 | 2.63·10<sup>4</sup> |
|  | PC2 | 0.02 | 2.58 | 0.15 | 0.00 | 3.8 | 0.00 | 3.45 | 0.12 | 0.00 | 4.00 | META | 2.20 | 0.50 | 0.115 | 0.83 | 5.88 |
| mLOY calls | mLOY | 1.14 | 0.14 | 0.35 | 0.87 | 1.49 | 1.04 | 0.16 | 0.81 | 0.76 | 1.43 | 65-70 | 0.55 | 0.45 | 0.19 | 0.22 | 1.30 |
|  | Age | 1.16 | 0.01 | 1.11·10<sup>-106</sup> | 1.15 | 1.18 | 1.28 | 0.01 | 1.73·10<sup>-71</sup> | 1.24 | 1.31 | 70-75 | 1.17 | 0.3 | 0.6 | 0.65 | 2.11 |
|  | APOE | 2.52 | 0.08 | 2.17·10<sup>-32</sup> | 2.17 | 2.94 | 2.86 | 0.10 | 6.93·10<sup>-28</sup> | 2.37 | 3.46 | 75-80 | 1.00 | 0.25 | 0.99 | 0.62 | 1.66 |
|  | PC1 | 2.29·10<sup>3</sup> | 2.59 | 2.78·10<sup>-3</sup> | 14.5 | 3.69·10<sup>-5</sup> | 0.00 | 3.32 | 0.10 | 0.00 | 2.96 | 80-85 | 3.70 | 0.77 | 0.09 | 1.00 | 24.08 |
|  | PC2 | 0.02 | 2.58 | 0.14 | 0.00 | 3.44 | 0.00 | 3.44 | 0.11 | 0.00 | 3.34 | META | 1.03 | 0.17 | 0.86 | 0.74 | 1.45 |

In order to check for an effect of mLOY in risk of conversion to all-cause dementia and AD, we fitted Cox proportional-hazards models adjusted by age at sampling and *APOE* genotype in our prospective cohort of MCI males (N=400). The continuous mLRR-Y variable had a non-significant risk effect in MCI conversion to all-cause dementia (HR=1.93; p=0.10). The effect size increased when we calculated the model exclusively using conversion to AD but did not reach statistical significance (HR=2.05; p=0.19). mLOY calls also showed similar but smaller non-significant positive effects for conversion to dementia (HR=1.17; p=0.40) and AD (HR=1.38; p=0.20, Figure 2). Cox model results are summarized in Table 2.

**Figure 2.**
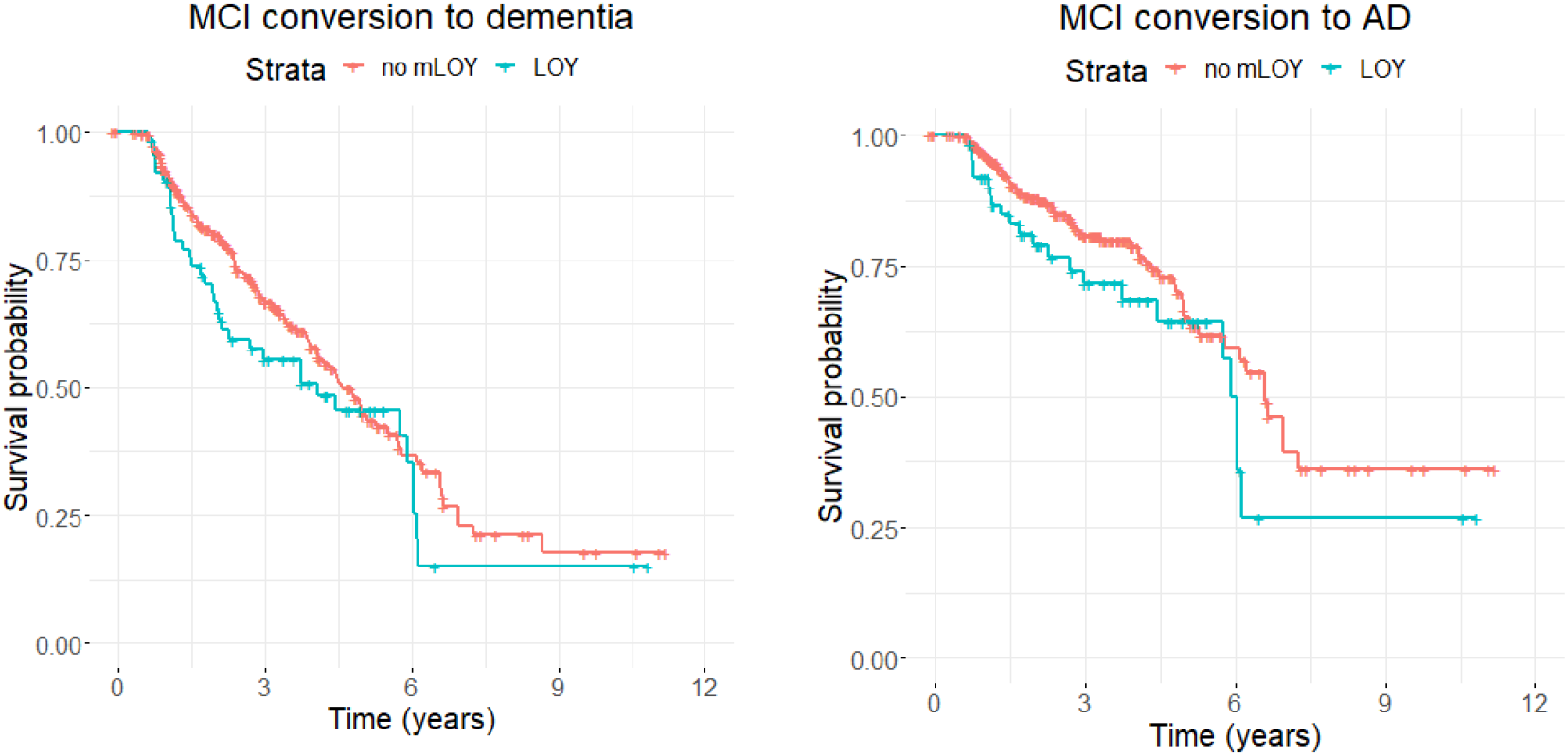
Association of mLOY phenotypes with risk of conversion to dementia and AD-dementia over time for MCI males in the GR@ACE-DEGESCO cohort. Kaplan-Meier plots showing survival time in years for conversion to dementia (**a**) or AD (**b**) for individuals with LOY (blue) or without LOY (red) for prospective MCI males in the GR@ACE-DEGESCO cohort.

**Table 2.** Cox proportional-hazards model results for conversion of MCI males to all-cause-dementia or AD-dementia. Models were adjusted by age at sampling and *APOE* genotype.

|  | Conversion to all-cause-dementia |  |  |  | Conversion to AD-dementia |  |  |  |
| --- | --- | --- | --- | --- | --- | --- | --- | --- |
|  | HR | SE | P-value | CI95 % | HR | SE | P-value | CI95 % |
| *Association results for mLRR-Y (continuous mLOY variable)* |  |  |  |  |  |  |  |  |
| **mLRR-Y** | 1.93 | 0.40 | 0.10 | 0.87-4.27 | 2.05 | 0.55 | 0.19 | 0.70-5.97 |
| **Age** | 1.10 | 0.01 | $3.62 \cdot 10^{-15}$ | 1.07-1.12 | 1.13 | 0.02 | $2.38 \cdot 10^{-12}$ | 1.09-1.16 |
| **APOE** | 1.29 | 0.13 | $4.10 \cdot 10^{-02}$ | 1.01-1.66 | 1.56 | 0.17 | $8.89 \cdot 10^{-3}$ | 1.12-2.17 |
| *Association results for mLOY calls* |  |  |  |  |  |  |  |  |
| **mLOY** | 1.17 | 0.19 | 0.40 | 0.81-1.70 | 1.38 | 0.25 | 0.20 | 0.85-2.24 |
| **Age** | 1.10 | 0.01 | $1.33 \cdot 10^{-15}$ | 1.07-1.13 | 1.13 | 0.02 | $1.62 \cdot 10^{-12}$ | 1.09-1.17 |
| **APOE** | 1.28 | 0.13 | $5.09 \cdot 10^{-2}$ | 1.00-1.63 | 1.54 | 0.17 | $1.05 \cdot 10^{-2}$ | 1.11-2.13 |

Because the impact of age on mLOY and AD might obscure genuine associations between both phenotypes (mLOY and AD), we decided to construct a mLOY Polygenic Risk Score (mloy-PRS) to evaluate the impact of the genetic variance associated with mLOY phenotype in AD risk. Our rationale was to implement a Mendelian randomization strategy reasoning that, if blood cell mLOY is genuinely associated with AD, the genetic factors linked to blood mLOY risk should be also associated with AD and its related endophenotypes. To this end, mloy-PRS was generated based on a list of autosomal genome-wide significant SNPs associated with the mLOY phenotype identified in a recent genome-wide association study (Supplementary Table 2) [14].

For benchmarking purposes of the constructed PRS, we initially validated the effect of mloy-PRS in the mLOY cell phenotype in our cohort (65–85 years old). To this end, we fitted a logistic regression for mLOY calls with PRS, age at DNA sampling and *APOE* genotype as predictors. The PRS (OR=1.80; p=4.22·10^−20^) and age at sampling (OR=1.08; p=5.07·10^−11^), but not *APOE* (OR=0.88; p=0.15), were significantly associated with mLOY in our population (Supplementary Table 7). Importantly, the PRS was orthogonal with age and evenly distributed across the age spectrum (Supplementary Figure 7). Our results reassure the validity of mloy-PRS as a mendelian randomization instrument for investigating the causal role of mLOY in AD and its endophenotypes, and independently confirm the combined risk effect of previously reported loci in the mLOY phenotype [14].

In the case-control setup, we checked the effect of mloy-PRS on AD risk by fitting logistic regressions adjusted by *APOE*, age and relevant PCs (Supplementary Table 8). Interestingly, the effect of mloy-PRS on AD could be measured in the female samples as well. Therefore, 3 analysis groups were stablished: all (males + females) and stratified by sex. No association between mloy-PRS and AD was found in the group including both sexes (Table 3). However, after sex stratification, we found a weak non-significant positive effect of mloy-PRS with respect to AD in the male subgroup (N=2 471; OR=1.07; p=0.12), while the effect was mostly neutral in the female subset (N=4 978; OR=1.00; p=0.93). Next, we assessed the effect of mloy-PRS in disease progression. We adjusted Cox models by age, *APOE* genotype and cohort ascertainment. We found a male-specific positive effect of mloy-PRS in the disease progression models (N=682) (Table 3, Supplementary Figure 8), with a suggestive signal for MCI to dementia progression (HR=1.11; p=0.08) and a statistically significant risk effect for MCI to AD progression (HR=1.23; p=0.01). Of note, no association between mloy-PRS and conversion to all-cause dementia (HR=0.99, p=0.81) or AD (HR=0.99; p=0.85) was found in the female group (N=1 082).

**Table 3.**
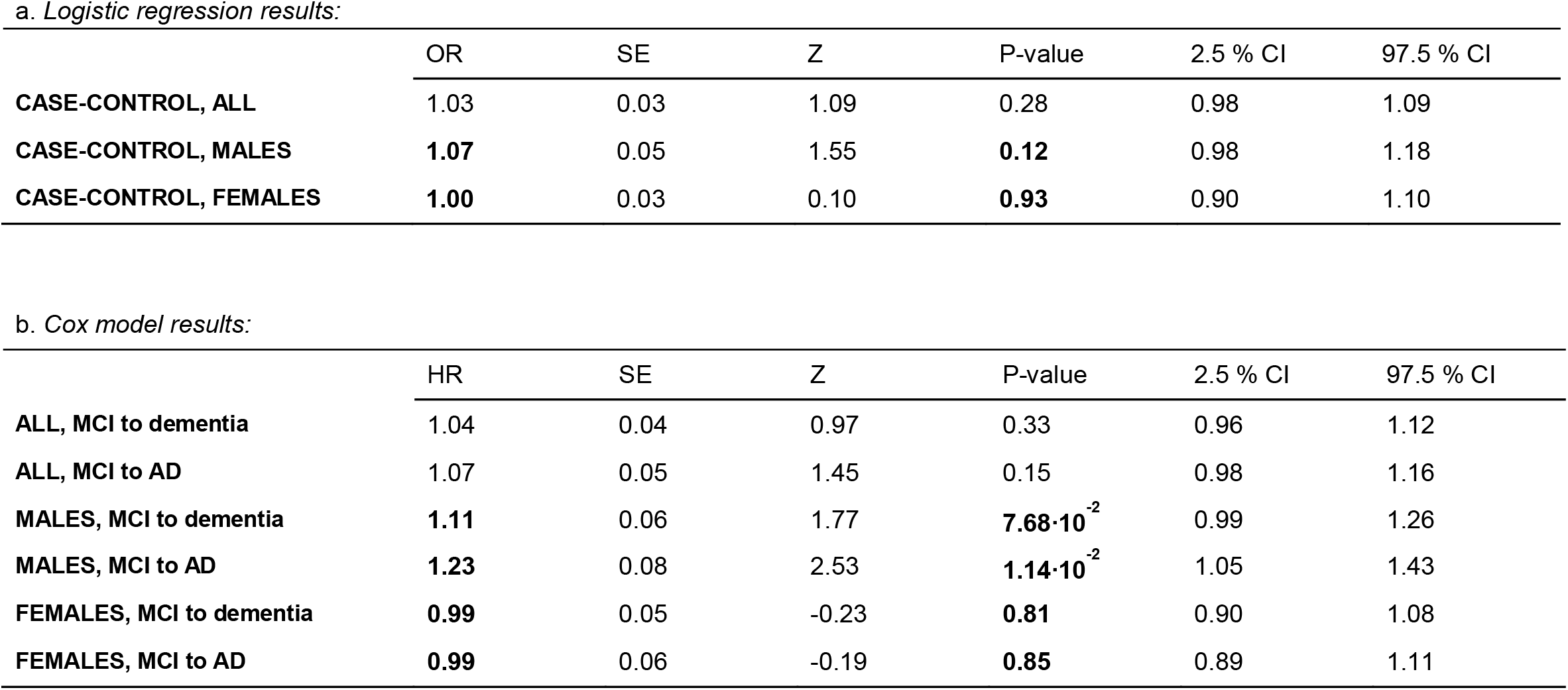
Association results for mLOY PRS. (**a)** Results of logistic regressions for case-control AD. Only individuals aged 65–85 were included in the models. Models were adjusted by *APOE*, age, and principal components. (**b**) Results of joint analysis of prospective MCIs in the GR@ACE-DEGESCO and EADB-DEGESCO cohorts using Cox proportional-hazards models for progression from MCI to all-cause dementia or AD. Models were adjusted by APOE, age, and cohort ascertainment.

Following these results, we proceeded to examine the existence of associations between mLRR-Y, the continuous mLOY variable, and the levels of core AD biomarkers in CSF (Abeta-42, p-tau and total tau). Only individuals aged 65–85 at the moment of lumbar puncture (LP) were kept, and those with a gap higher than 5 years between DNA sampling and the LP were excluded from the analysis (N=214). Linear regressions were adjusted by *APOE* genotype, age at LP, and time window between blood sampling and LP. To account for the effect of syndromic status on the levels of Abeta-42, p-tau and total tau, we calculated the effect in two groups (MCI N=148; dementia N=66) and then performed inverse-variance weighted fixed effect meta-analysis of the effect of mLRR-Y. We found that both p-tau (β=41.92; p=0.01) and total tau (β=396.69; p=0.004) levels were increased in individuals with a higher degree of mLOY (Figure 3).

**Figure 3.**
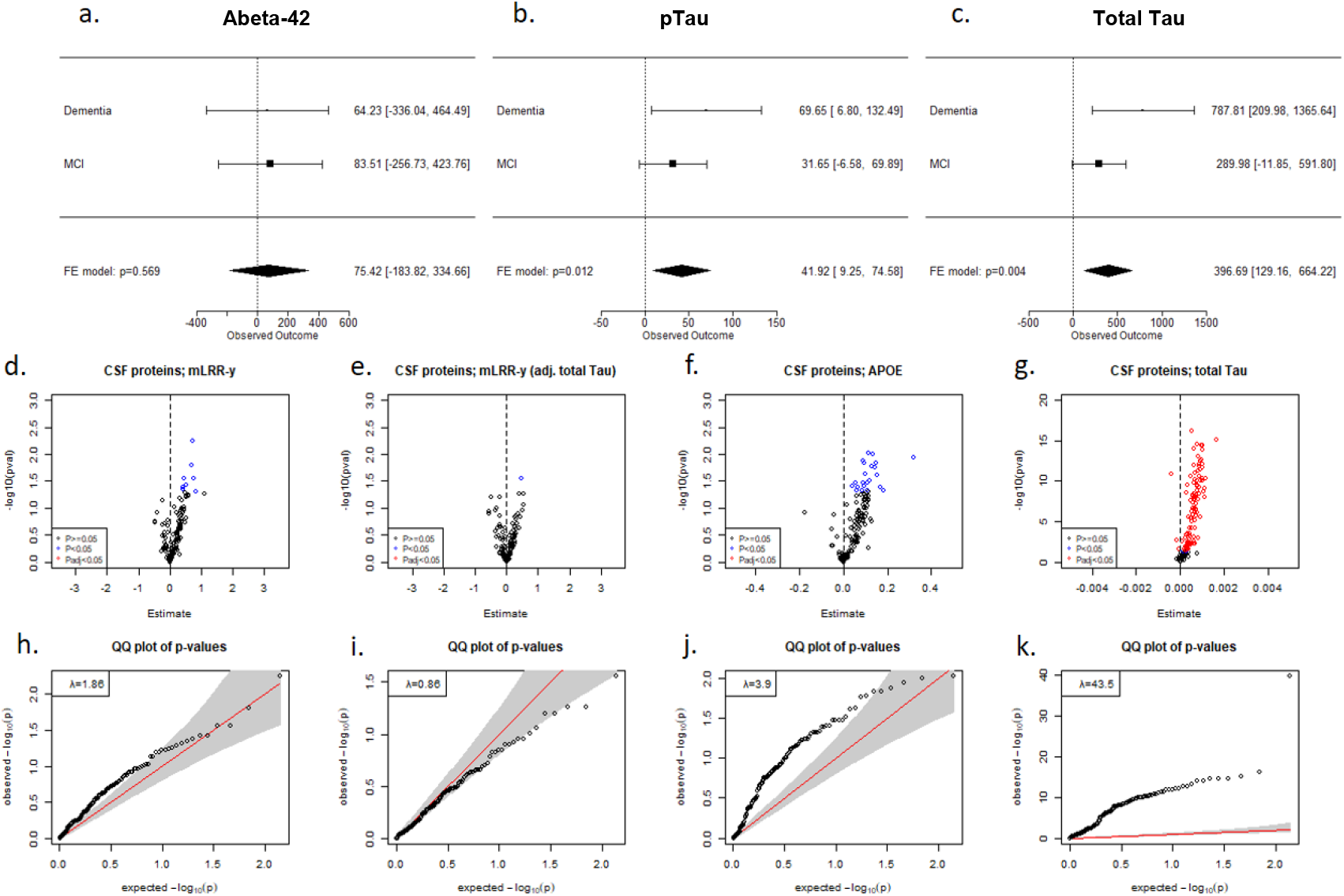
mLRR-Y associations with CSF protein levels. **a-c**. Forest plots showing the effect size obtained in linear regression models for mLRR-Y on Abeta-42 (**a**), phospho-Tau (**b**), and total Tau (**c**) in males with MCI or dementia, along with meta-analysis results. **d–g**. Volcano plots showing association of CSF proteins in the Olink Inflammation and Neurology panels with mLRR-Y (**d**), mLRR-Y adjusted by total Tau (**e**), *APOE* genotype (**f**), and total Tau (**g**). **h– k**. QQ plots obtained in the models for mLRR-Y (**h**), mLRR-Y adjusted by total Tau (**i**), *APOE* genotype (**j**), and total Tau (**k**). Models were adjusted by age, the time window between CSF and DNA sampling, and *APOE* genotype.

Afterwards, we checked for mLRR-Y associations with proteomics data obtained with the Olink ProSeek® multiplex immunoassay for paired plasma and CSF samples in 135 MCI males. Because mLOY is known to affect the immune system [10, 24, 54], and inflammation is involved in many processes related to AD pathogenesis [55], we analysed Olink Neurology and Inflammation panels. We detected inflation in our models (λ=1.86), with most proteins showing increased levels in the CSF of individuals with a higher degree of blood mLOY (Figure 3). A similar pattern was observed when we analysed the effect of *APOE* genotype and total tau levels, with a big fraction of the proteins showing increased CSF levels in individuals carrying *APOE* risk alleles or displaying higher tau levels, respectively (Figure 3). Moreover, after adjusting our models by total tau we lost most CSF associations and the inflation factor was drastically reduced to λ=0.86 (Figure 3). After covariation with total tau, we found 7 nominally significant markers in plasma and 1 nominally significant marker in CSF. Nevertheless, no proteins passed FDR correction suggesting that most mLOY associations can be explained by the previously observed correlation between mLOY and tau levels. Summary statistics for association of mLRR-Y to the CSF and plasma proteins are available (Supplementary Tables 9-12).

## DISCUSSION

In the present study, we found that MCI males with high genetic risk of developing mLOY have increased chances of progressing to AD over time. The autosomal loci used to construct the mLOY PRS had no effect on AD progression in the female subset of our cohort, strongly suggesting that the observed effect is produced via loss of the Y chromosome among men. Importantly, modelling mLOY through its associated genetic variance allowed us to observe mLOY-induced alterations in AD pathogenesis in an age-independent manner, unparalleled in previous studies. These results add to previous evidence reporting mLOY as a male-specific AD pathogenic factor.

mLOY is the most common known form of somatic mosaicism among men [10]. Concordantly, we detected mLOY in 18.9% men aged 65–85 in our cohort. Though classically thought of as a harmless age-related trait, recent studies have revealed that mLOY increases risk for all-cause mortality and several diseases [10, 20–22]. With such a high prevalence in the elderly population, interest in determining mLOY’s effect in age-related diseases has raised over the past decade. Previous studies reported that mLOY significantly increases AD risk and progression rate [19]. A more recent publication claims that transcriptomic extreme downregulation of chromosome Y decreases AD resilience in men [56]. However, whether mLOY acts as an AD promoting factor or is just a by-product of ageing is not yet clearly established.

Concordant with previous studies [19], we found a higher degree of mLOY mosaicism (mLRR-Y) in our AD vs. control population in unadjusted Kolmogorov-Smirnoff models (D=0.18124; p < 2.2e-16) and age-adjusted ANCOVA (F=68.0; p=2.54·10^−16^). We then performed a case-control logistic regression in all available males in our cohort, obtaining statistically significant results (OR=2.74; p=0.01). However, even though age was adjusted in our data, these results should be taken cautiously due to the dramatic age differences between the AD and control groups (Figure 1), as age is an important risk factor for both phenotypes. Thus, aiming to reduce age confounding in our models, we restricted analysis to men aged 65–85 years old. However, despite not completely correcting the age gap between the groups, this also reduced our sample size (Supplementary Table 4). We found an increased degree of LOY mosaicism (mLRR-Y) in 65–85-year-old cases vs. controls (OR=2.19; p=0.09), but statistical significance was not reached in the models.

Previous reports also found that mLOY increases the rate of AD conversion in MCI males [19]. We selected individuals recruited in ACE Alzheimer Center with an MCI diagnosis at the moment of sampling and available clinical follow ups, and fitted Cox proportional-hazards models. Even though we found risk, or positive, effect directions for mLOY phenotypes towards AD progression, no statistical significance was reached in the models (Figure 3). However, given our small sample size (N=400), we may have lacked enough statistical power in this analysis. Remarkably, we noticed that the quantitative mLRR-Y variable performed superiorly in the case-control and disease progression models compared to mLOY calls, implying that, if effects were genuine, the mLOY-induced increase in AD risk and progression may be proportional to the mosaic fraction of LOY cells in blood.

Due to the age-dependent nature of both mLOY and AD, controlling age confounding was very challenging in our cohort. For this reason, we checked mLOY causality in AD by creating an instrument variable and conducted a mendelian randomization study. To this end, we generated an age-independent and sex-independent mLOY-associated PRS, using 114 independent autosomal genetic variants (Supplementary Table 2) previously associated with mLOY [14]. Of note, mloy-PRS successfully predicted mLOY events in our data and was not associated with age or *APOE* genotype (Supplementary Figure 7, Supplementary Table 7). A recently published work found similar effect sizes of this PRS for predicting mLOY calls [57]. Therefore, analysis of mloy-PRS instead of mLOY phenotypes allowed us to overcome the main limitations of the study (age differences and sample size) by providing an age-independent mLOY instrument, and allowing us to increase the effective sample size in two ways: (a) removing the need to restrict analysis to samples with available age at DNA sampling information and (b) allowing us to introduce all MCI individuals with subsequent clinical records in disease progression models instead of only those with an MCI diagnose at the closest clinical evaluation to DNA sampling.

Importantly, we found a male-specific, statistically significant (HR=1.23; p=0.01) association between mloy-PRS and MCI phenoconversion to AD. Case-control models also reported positive, or risk, effects of mloy-PRS in AD in the male subset (OR=1.07; p=0.12), but no statistical significance was reached in the models, even though our sample size was considerably larger in the case-control dataset (N_males_=2 471) than in the longitudinal, prospective MCI dataset (N_males_=682). These results suggest that mLOY could be more involved in the MCI, early clinical stages of AD etiopathogenesis than in the preclinical stages of the disease, namely AD risk. However, because the PRS only explains a fraction of the variance that causes mLOY, a higher sample size may be needed to reach enough statistical power to obtain more robust associations in the case-control models. Importantly, mloy-PRS effects were neutral in the female groups (Table 3), implying that the observed effect of mloy-PRS on AD is unlikely to be driven by the same mechanisms that confer mLOY risk (increased genomic instability and impairment of DNA reparation mechanisms) [14]. Instead, the observed effects were male-specific and therefore more likely produced via loss of the Y chromosome exclusively in men.

One of the most commonly proposed mechanisms to explain blood cell LOY pathogenesis is the impairment of immune functions [10, 14, 19]. Interestingly, deregulation of the immune system is one of the hallmark features of AD [58], and genome-wide association studies are revealing an increasing number of genes related to immune functions [30]. LOY has been reported to cause deregulation in the expression levels of approximately 500 autosomal transcripts in leukocytes [24]. Furthermore, levels of CD99, a cell surface protein involved in several key immune functions, such as leukocyte migration through the vascular endothelium, cell adhesion and apoptosis [59, 60], have been found to be significantly lowered in immune cells with LOY [24, 54]. Thus, mLOY-induced alterations in the homeostasis and migration of leukocytes through the brain-blood barrier could explain the observed associations. Further studies are necessary to corroborate our findings and to identify the specific mechanisms within mLOY modifying AD etiopathogenesis. Of note, functional restauration of the lost Y-chromosome loci promoting aberrant clonal expansion or transcriptomic deregulation of LOY leukocytes could be an attractive therapeutic strategy for combating AD progression.

One strength of our study is that we modelled LOY through an age-independent PRS instead of just analyzing the age-dependent mLOY phenotype and adjusting our data by age. In our opinion, this allowed a clearer and more robust approach for inferring causality between mLOY and AD. We also obtained an independent validation of our findings through AD-related biomarkers, with mLOY phenotypes associated with higher levels of total tau and p-tau in the CSF and displaying the proteomic neurodegenerative biochemical signature observed with other AD-related factors (Figure 3). Higher tau levels are associated with faster rates of cognitive decline [61], supporting the hypothesis that mLOY modulates disease progression. This work, however, also faced several limitations: a) the lack of age at sampling information for most controls and b) the control population being significantly younger than the AD population. Both of these limitations ultimately decreased our statistical power to find more robust mLOY-AD associations in the case-control models.

In summary, the present study did not find such strong associations between the blood mLOY phenotype and AD as those reported previously [19]. Due to the demographic features of the GR@ACE/DEGESCO cohort, with older AD patients and younger population-based controls, adjusting our data by age was challenging. Consequently, we modelled the genetic variance associated with mLOY risk, generating a PRS that was associated with MCI conversion to AD in a male-specific manner. This approach allowed us to efficiently control the effect of ageing and to evaluate the potential causality of the mLOY phenotype. Furthermore, lack of association of mloy-PRS and AD in women suggests that the observed effect is produced via the inherent loss of the Y chromosome and that mLOY could be a male-specific AD risk factor. Larger studies may benefit from modelling mLOY using Mendelian randomization, as case and control populations do not always represent the same age groups in AD cohorts, and date of DNA sampling of the subjects may not be available.

## Data Availability

All data produced in the present study are available upon reasonable request to the authors.

## AKNOWLEDGEMENTS

We would like to thank patients and controls who participated in this project. The present work has been performed as part of the doctoral thesis of P.G. at the University of Barcelona (Barcelona, Spain). P.G. is supported by CIBERNED employment plan CNV-304-PRF-866. CIBERNED is integrated into ISCIII (Instituto de Salud Carlos III). I. de Rojas is supported by a national grant from the Instituto de Salud Carlos III FI20/00215. A. Cano acknowledges the support of the Spanish Ministry of Science, Innovation and Universities under the grant Juan de la Cierva (FJC2018-036012-I). M.B. and A.R. are also supported by national grants PI13/02434, PI16/01861, PI17/01474, PI19/01240 and PI19/01301. The Genome Research @ Fundacio ACE project (GR@ACE) is supported by Grifols SA, Fundación bancaria La Caixa, Fundació ACE, and CIBERNED. Acción Estratégica en Salud is integrated into the Spanish National R□+□D□+□I Plan and funded by ISCIII (Instituto de Salud Carlos III)—Subdirección General de Evaluación and the Fondo Europeo de Desarrollo Regional (FEDER—Una manera de hacer Europa). Some control samples and data from patients included in this study were provided in part by the National DNA Bank Carlos III (www.bancoadn.org, University of Salamanca, Spain) and Hospital Universitario Virgen de Valme (Sevilla, Spain); they were processed following standard operating procedures with the appropriate approval of the Ethical and Scientific Committee. Genotyping of the ACE MCI-EADB samples was performed in the context of EADB (European Alzheimer DNA biobank) funded by the JPco-fuND FP-829-029 (ZonMW project number 733051061). This work was supported by a grant (European Alzheimer DNA BioBank, EADB) from the EU Joint Program—Neurodegenerative Disease Research (JPND).

## Notes

### Competing Interest Statement

The authors have declared no competing interest.

### Author Declarations

Written informed consent was obtained from all participants. The Ethics and Scientific Committees have approved this research protocol (Acta 25/2016, Ethics Committee. H., Clinic I Provincial, Barcelona, Spain)

