## Supplementary Figure 1 for "Mendelian randomization confirms the role of Y-chromosome loss in Alzheimer’s Disease etiopathogenesis in males"

**Supplementary Figure 1. Overview of mLRR-Y and PAR1-Bdev distributions in our dataset. a**. Plot showing mLRR-Y and pseudoautosomal región 1 B-deviation (PAR1-Bdev) values for male samples in the GR@ACE/DEGESCO cohort. Red-colored dots represent samples displaying anomalies, which were discarded from further analysis. After exploring PAR1 BAF probe distributions in our dataset, we considered heterozygous probes as those with BAF values in the range of 0.23–0.77. mLRR-Y and PAR1-Bdev were highly correlated ($R2=0.92$; $p<2.2\cdot10^{-16}$) in samples with low to moderate degrees of LOY mosaicism (mLRR-Y ranging -0.1 to -0.6). However, for samples with a higher proportion of LOY in blood cells (mLRR-Y<-0.6), BAF values of heterozygous probes became increasingly distant from 0.5 and were not efficiently captured using fixed thresholds, affecting the method accuracy for obtaining PAR1-Bdev in samples with mLRR-Y<-0.6 and dramatically reducing PAR1-Bdev correlation with mLRR-Y ($R2=0.08$; $p=0.01$) within these samples. Thus, capturing heterozygous probes for samples with a high degree of mLOY would require using a wider BAF window. This phenomenon was clearly evidenced when we observed PAR1 B allele frequency (PAR1 BAF) distributions of samples with no LOY (**b**), moderate mLOY (**c**), or extreme mLOY (**d**).

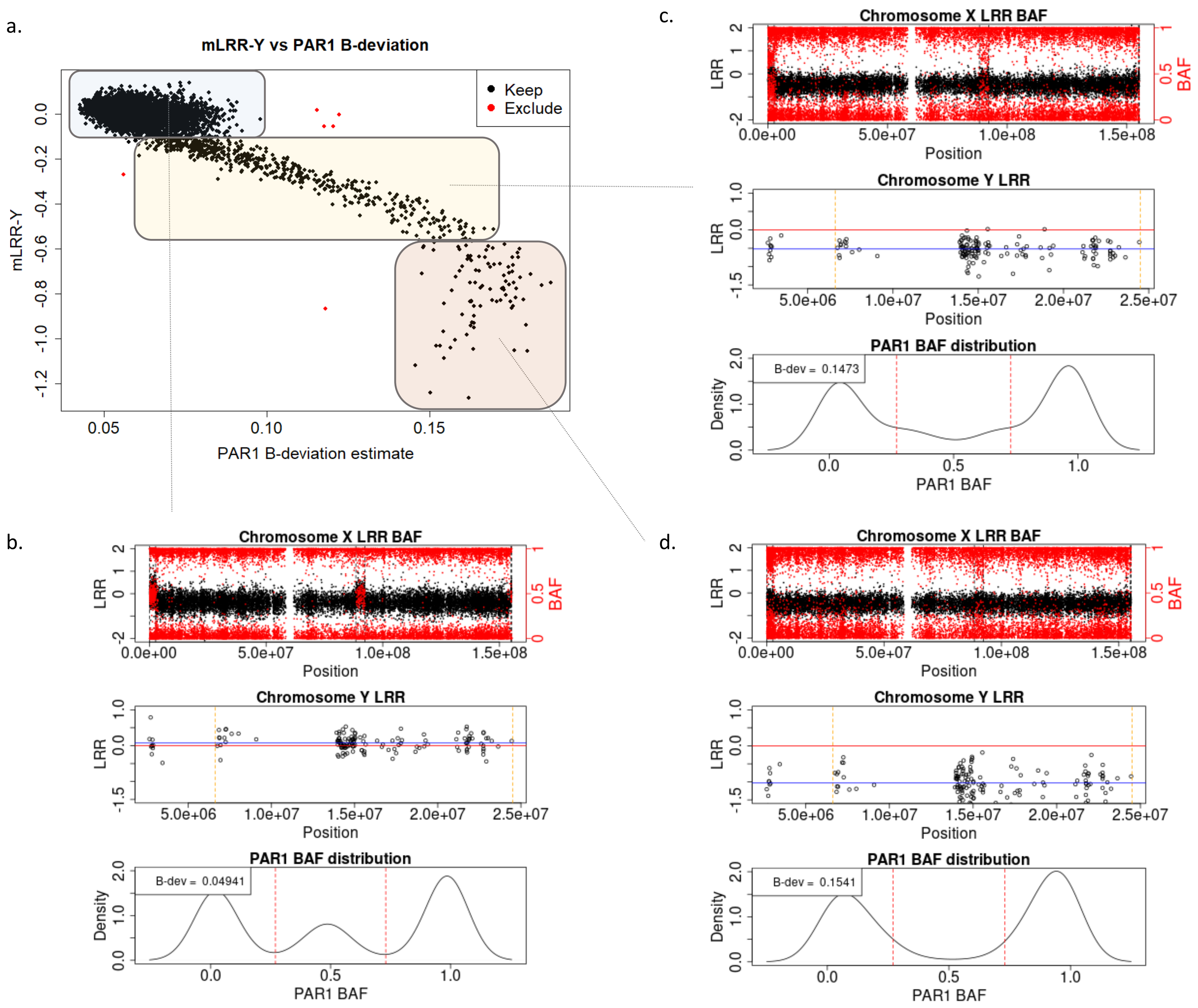
