## Supplementary Figure 2 for "Mendelian randomization confirms the role of Y-chromosome loss in Alzheimer’s Disease etiopathogenesis in males"

**Supplementary Figure 2**. Flow chart describing QC and filtering steps prior to case-control and disease progression analysis, depending on the use of mLOY phenotype or mloy-PRS as the independent variable.

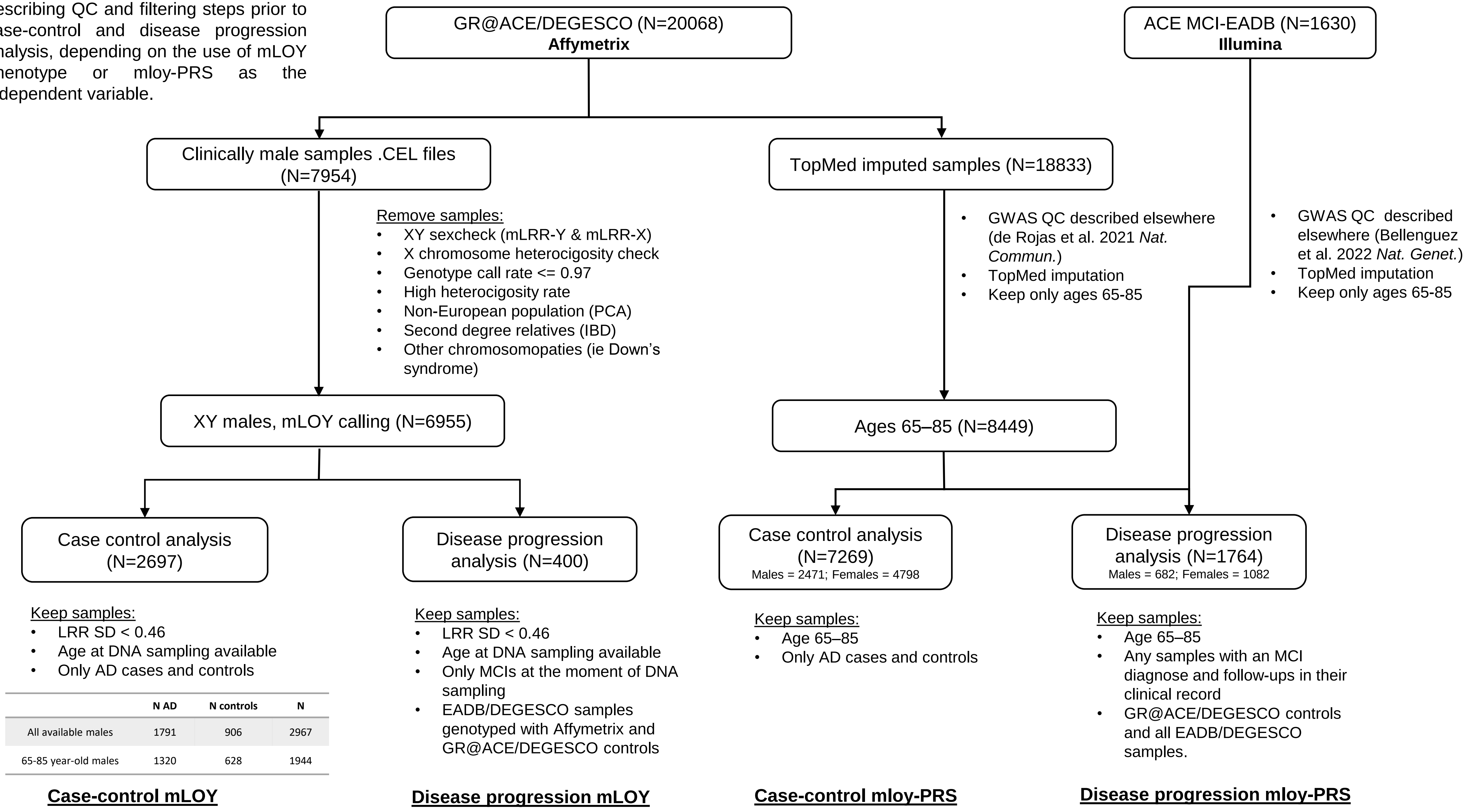


| | N AD | N controls | N |
|---|---|---|---|
| All available males | 1791 | 906 | 2967 |
| 65-85 year-old males | 1320 | 628 | 1944 |
