## Supplementary Figure 3 for "Mendelian randomization confirms the role of Y-chromosome loss in Alzheimer’s Disease etiopathogenesis in males"

**Supplementary Figure 3. PCs asociation to conversión from MCI to dementia (PRS analysis).** PC1 is highly associated to AD, but in fact represents genotyping chip/cohort ascertainment as shown. Thus, PC1 was not included in the models, as we already adjusted our data by genotyping chip/cohort ascertainment.

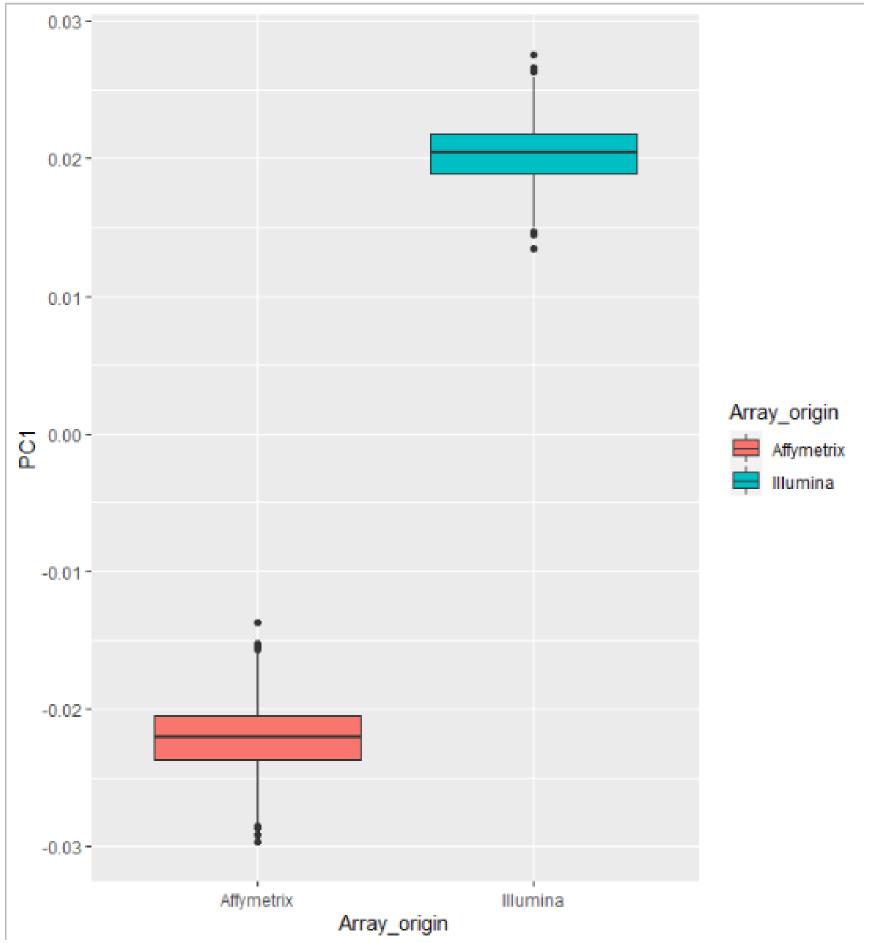


| | coef | exp(coef) | se(coef) | z | Pr(>\|z\|) | 2.5 % | 97.5 % |
|---|---|---|---|---|---|---|---|
| **PC1** | -18.58 | $8.50 \cdot 10^{-9}$ | 1.63 | -11.38 | **5.45**$\cdot 10^{-30}$ | -21.78 | -15.38 |
| PC2 | 1.44 | 4.21 | 1.54 | 0.94 | 0.35 | -1.57 | 4.45 |
| PC3 | -0.21 | 0.81 | 1.58 | -0.13 | 0.90 | -3.29 | 2.88 |
| PC4 | -1.60 | 0.20 | 1.58 | -1.01 | 0.31 | -4.69 | 1.49 |
| PC5 | -1.97 | 0.14 | 1.60 | -1.23 | 0.22 | -5.10 | 1.16 |
