## Supplementary Figure 4 for "Mendelian randomization confirms the role of Y-chromosome loss in Alzheimer’s Disease etiopathogenesis in males"

**Supplementary Figure 4**. Heatmap displaying Pearson correlation coefficients (R) for total tau, p-tau and CSF proteins measured with Olink Neurology (a) and Inflammation (b) panels. Olink NPX values were normalized (mean=0, SD=1) prior to computing R coefficients.

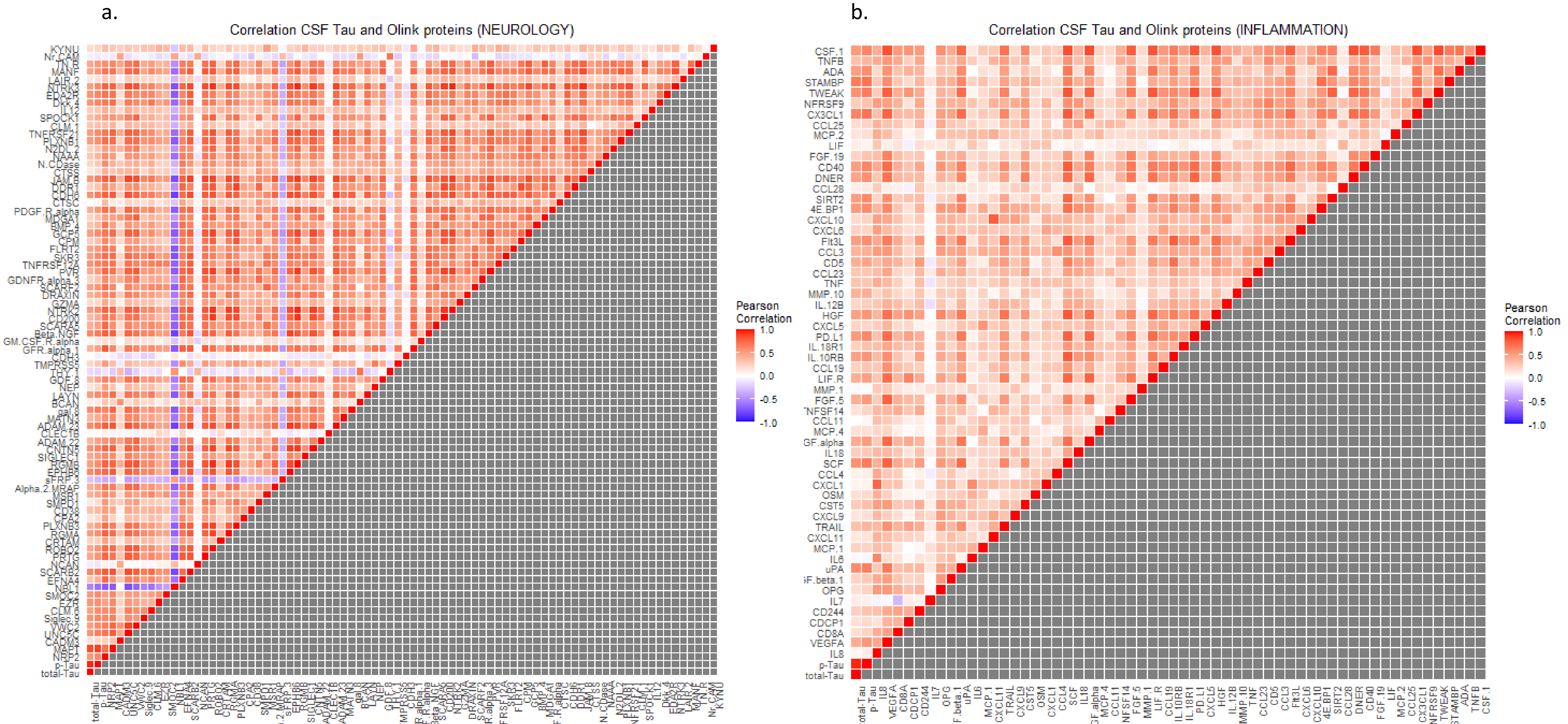
