## Supplementary Figure 5 for "Mendelian randomization confirms the role of Y-chromosome loss in Alzheimer’s Disease etiopathogenesis in males"

**Supplementary Figure 5. a**. Plot of mean log R Ratio for chromosome X (mLRR-X) and chromosome Y (mLRR-Y). The top left corner represents genetically normal XY men, with an observable degradation of mLRR-y values representing individuals with varying degrees of mLOY. Bottom right corner represents genetically normal women (XX), and top right corner Klineffelter (XXY) individuals. A GOY sample (XYY) was also identified above the top left cluster. **b-d.** identification and characterization of outliers of the PAR1 Bdev – mLRRy plot by representation of LRR and B allele frecuency (BAF) values.

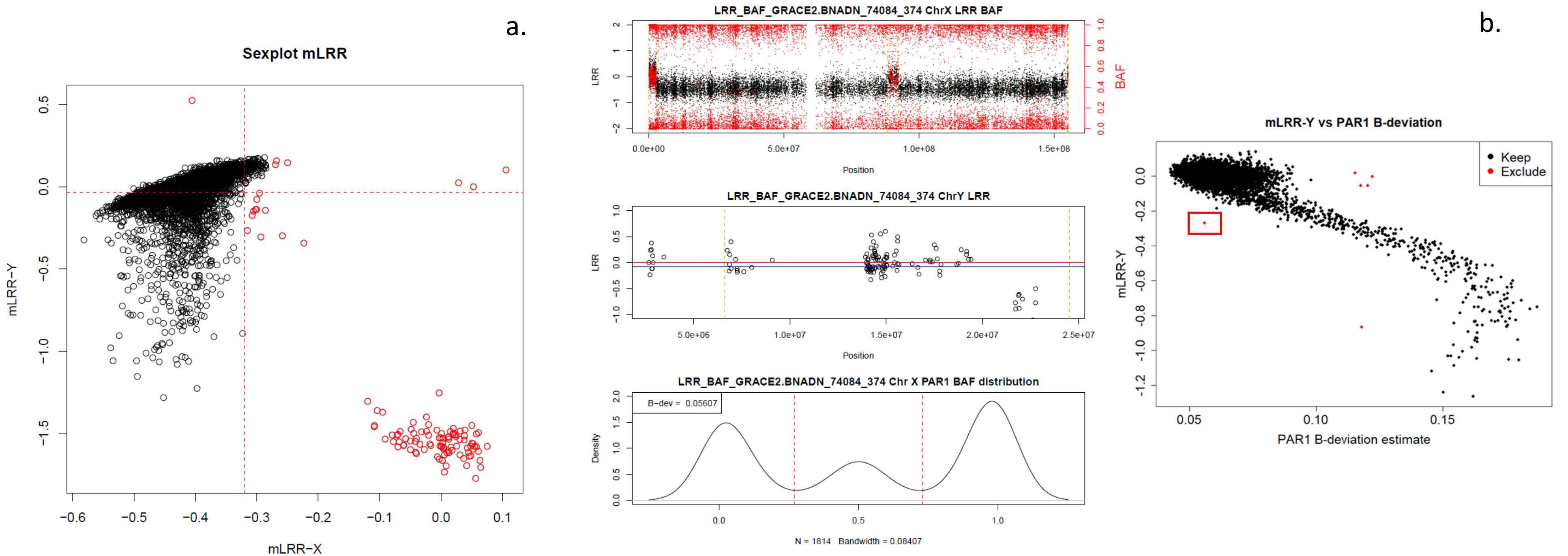


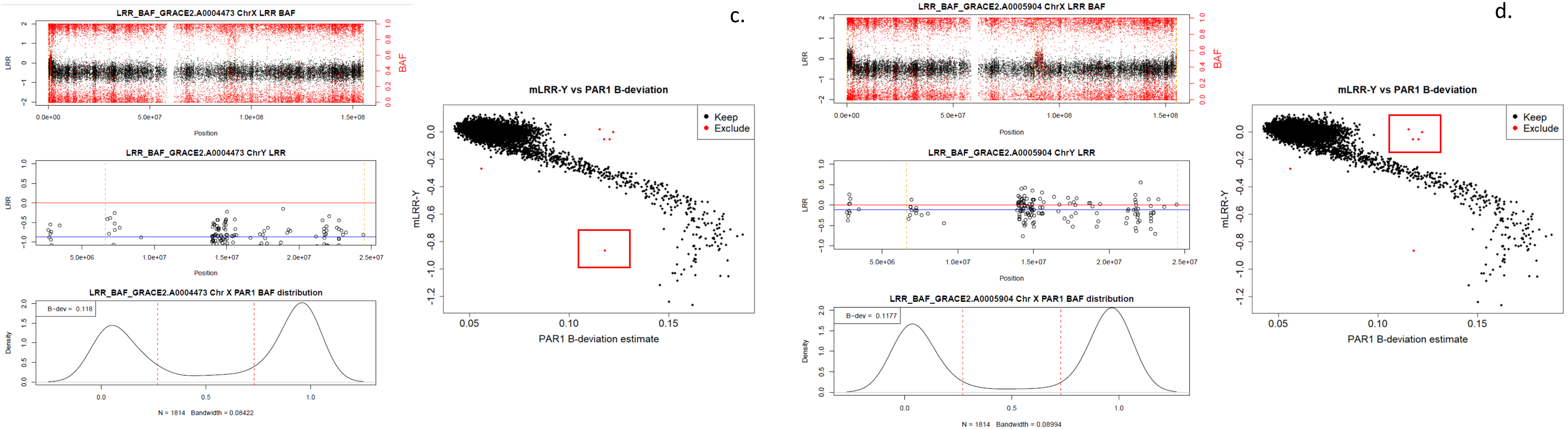
