## Supplementary Figure 6 for "Mendelian randomization confirms the role of Y-chromosome loss in Alzheimer’s Disease etiopathogenesis in males"

**Supplementary Figure 6. Análisis of batch effect in mLOY calling.** We tested a linear model using batch and age at extraction as predictors of mLRR-Y (continous mLOY parameter).

| | Beta | SE | P |
|---|---|---|---|
| **Batch** | $-6.02 \cdot 10^{-3}$ | $5.17 \cdot 10^{-3}$ | 0.24 |
| **Age** | $3.57 \cdot 10^{-3}$ | $2.75 \cdot 10^{-4}$ | $1.39 \cdot 10^{-37}$ |

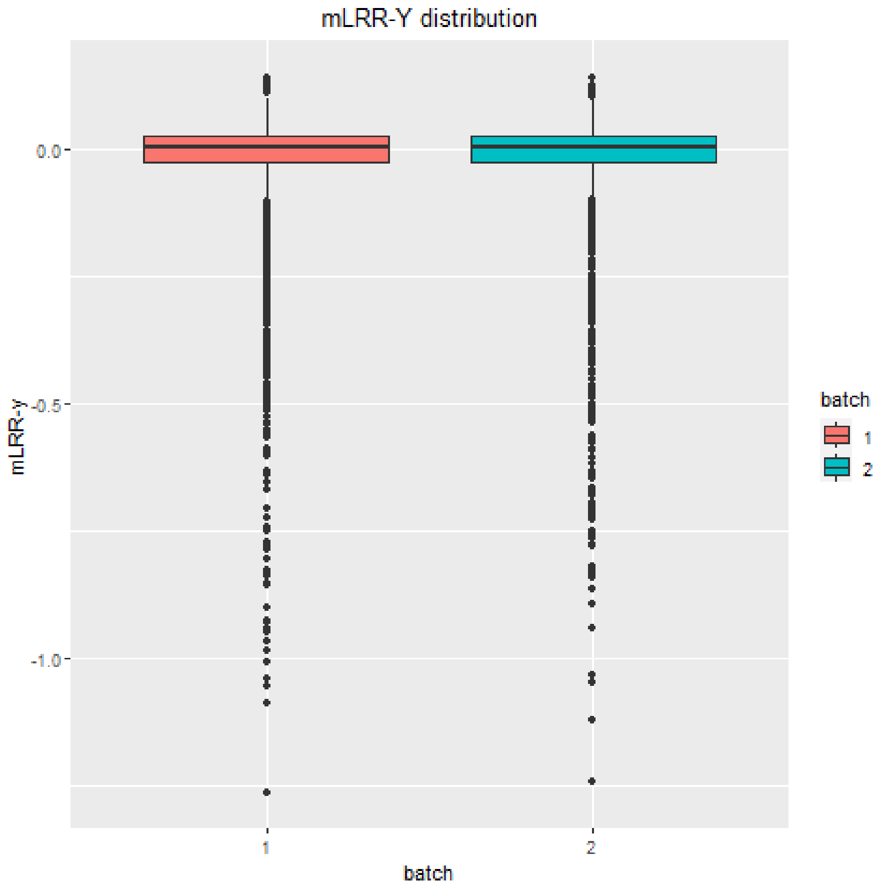
