## Supplementary Figure 7 for "Mendelian randomization confirms the role of Y-chromosome loss in Alzheimer’s Disease etiopathogenesis in males"

**Supplementary Figure 7. Validation and age Independence of the mloy-PRS variable. a.** PRS tertile distribution across the age spectrum. Colored lines represent the linear regression beta coefficients of mLRR-Y (dependent variable) and age (independent variable) for individuals classified in tertiles based on mloy-PRS. **b.** Linear regression of age (independent variable) with mLOY PRS (dependent variable) in the 65-85 age group.

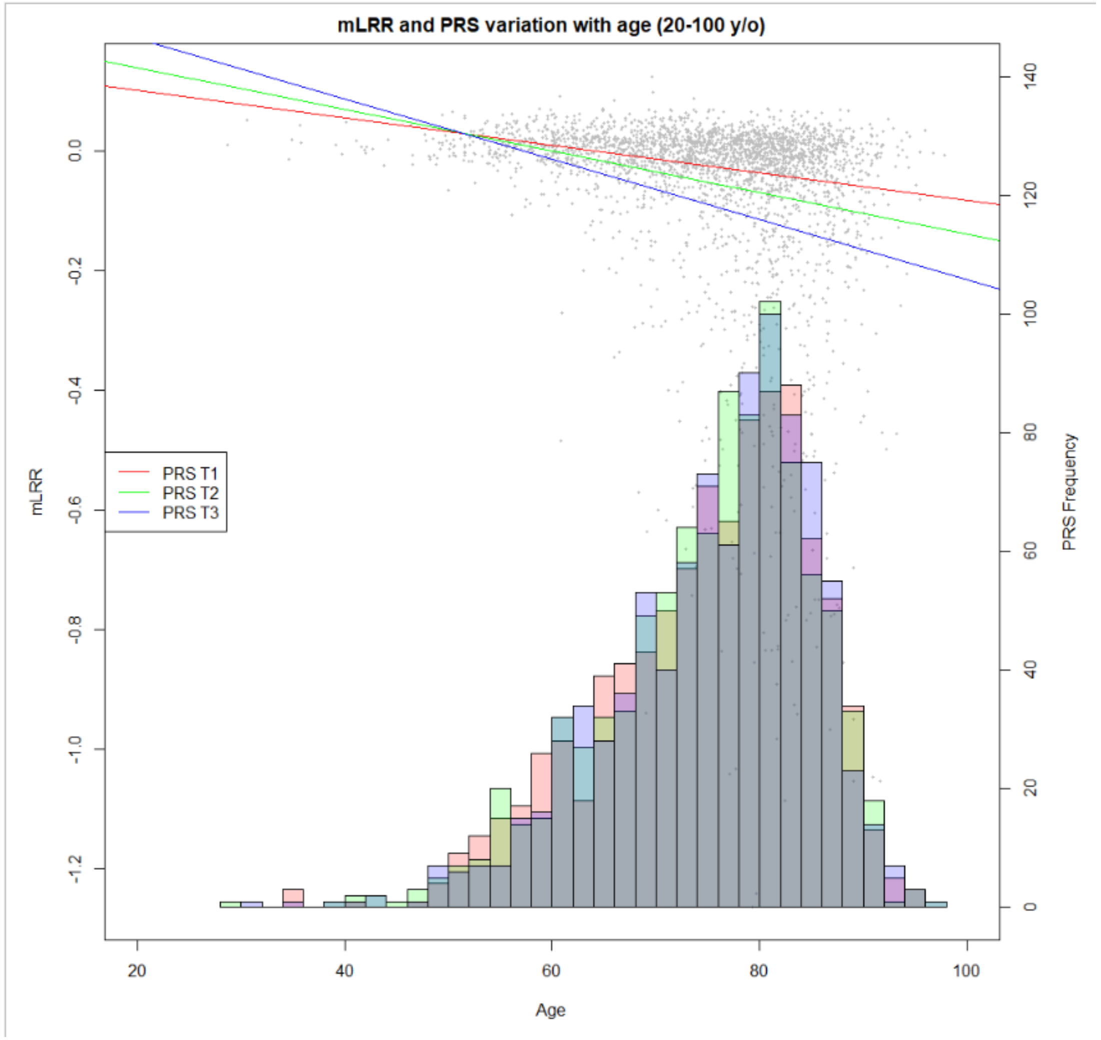


| b. | Beta | SE | t | P | CI2.5 | CI97.5 |
|---|---|---|---|---|---|---|
| **Age** | 0.00 | 0.02 | 0.13 | 0.90 | -0.03 | 0.04 |
