## Supplementary Figure 8 for "Mendelian randomization confirms the role of Y-chromosome loss in Alzheimer’s Disease etiopathogenesis in males"

**Supplementary Figure 8. mloy-PRS association with MCI progression to all-cause-dementia and AD-dementia.** Kaplan-Meier plots represent survival curves for individuals classified by mloy-PRS tertiles. Forest plots show the effect sizes of mloy-PRS, age, and *APOE* genotype in the models for conversion to dementia (**a**) or AD (**b**) in females and conversion to dementia (**c**) or AD (**d**) in males.

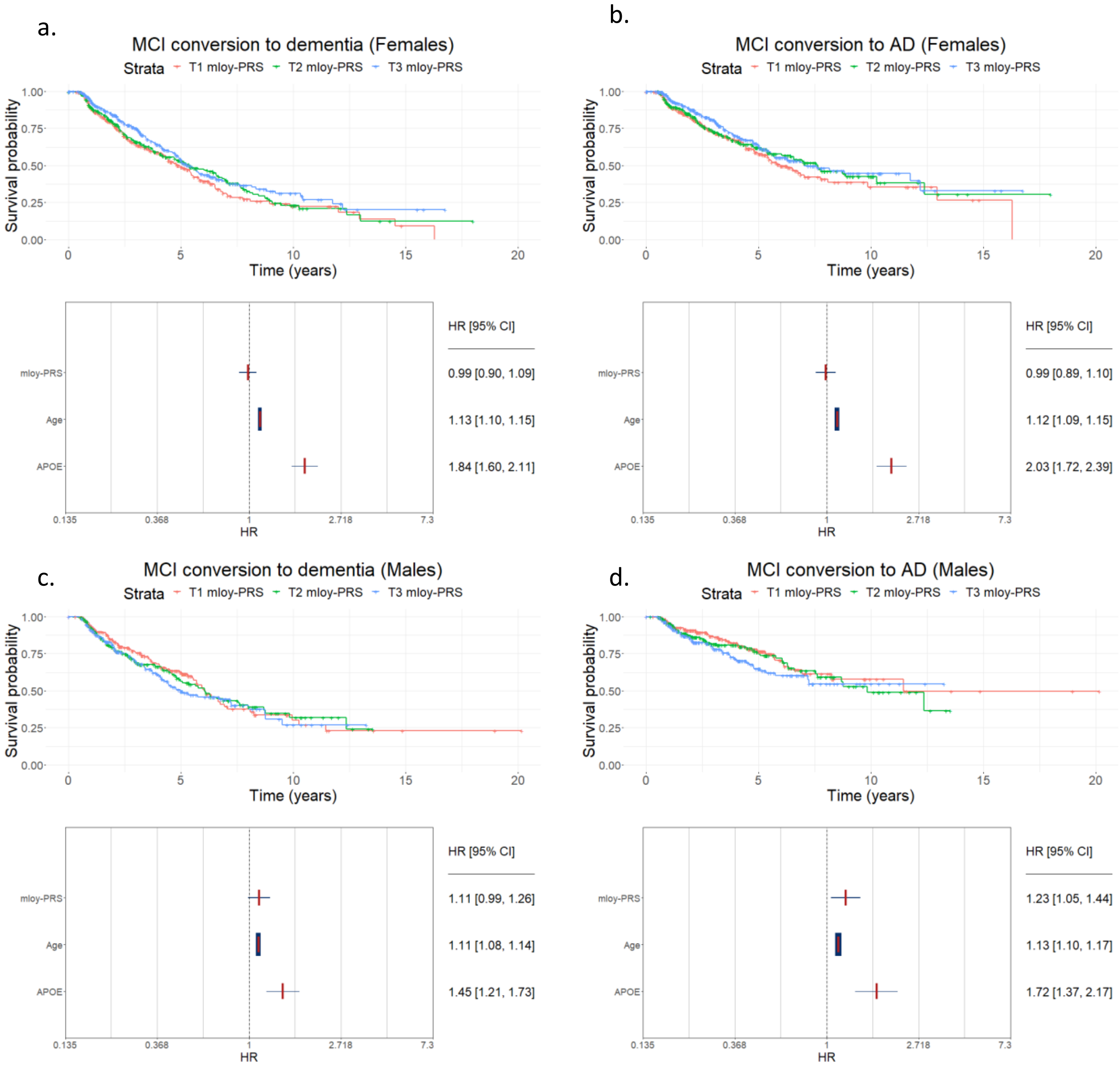
