## Supplementary Table 1 for "Mendelian randomization confirms the role of Y-chromosome loss in Alzheimer’s Disease etiopathogenesis in males"

**Supplementary Table 1. Sample contribution by center to the complete GR@ACE/DEGESCO cohort and different subanalysis selections.**

| Center | Source of DNA | GR@ACE/DEGESCO | PRS case-control | mLOY case-control | mLOY case-control (age 65-85) |
|---|---|---|---|---|---|
| ACE Alzheimer Center Barcelona | Blood | 8722 | 4926 | 2286 | 1552 |
| Banco Nacional de ADN | Blood | 3738 | 307 | | |
| Centro de Biología Molecular Severo Ochoa | Blood | 23 | 21 | | |
| Fundación CIEN | Blood | 1322 | 904 | 411 | 392 |
| HRC | Blood | 38 | 25 | | |
| Hospital Universitario Central de Asturias | Blood | 195 | 148 | | |
| Hospital Universitario de Valme | Blood | 1022 | 30 | | |
| IBIS | Blood | 1005 | 445 | | |
| IRBLleida | Blood | 230 | 160 | | |
| Mutua de Terrassa | Blood | 247 | 153 | | |
| Universidad de Málaga | Saliva | 408 | 78 | | |
| Hospital Universitario de Marqués de Valdecilla | Blood | 298 | 72 | | |
| | | ***INCLUSION CRITERIA:*** | • Age 65–85 | • Age at DNA sampling available<br>• Males | • Age at DNA sampling 65–85<br>• Males |
