## Supplementary Table 3 for "Mendelian randomization confirms the role of Y-chromosome loss in Alzheimer’s Disease etiopathogenesis in males"

**Supplementary Table 3. Cox Proportional-Hazards model results for PCs association to conversion from MCI to dementia in prospective MCIs from the GRACE-DEGESCO cohort.**

| | log(HR) | HR | SE | Z | P | 2.5 % | 97.5 % |
|---|---|---|---|---|---|---|---|
| PC1 | -4.28 | 0.01 | 3.92 | -1.09 | 0.27 | -11.96 | 3.39 |
| PC2 | -1.38 | 0.25 | 1.46 | -0.94 | 0.35 | -4.24 | 1.49 |
| PC3 | -0.48 | 0.62 | 1.65 | -0.29 | 0.77 | -3.72 | 2.76 |
| PC4 | -0.36 | 0.70 | 1.48 | -0.24 | 0.81 | -3.25 | 2.54 |
