## Supplementary Table 4 for "Mendelian randomization confirms the role of Y-chromosome loss in Alzheimer’s Disease etiopathogenesis in males"

**Supplementary Table 4. Age and sample size of the case and control populations in the GR@ACE-DEGESCO cohort before and after age filtering.** Only samples with age at DNA extraction available were considered.

| | N total | N group | Age total; Mean (SD) | Age groups; Mean (SD) | |
|---|---|---|---|---|---|
| All cases and controls | 2697 | 906 | 75.5 (9.5) | 68.7 (9.1) | Control |
| | | 1791 | | 79.0 (7.7) | AD |
| Age 65–85 | 1944 | 624 | 76.4 (5.2) | 73.5 (4.6) | Control |
| | | 1320 | | 77.8 (4.8) | AD |
