## Supplementary Table 5 for "Mendelian randomization confirms the role of Y-chromosome loss in Alzheimer’s Disease etiopathogenesis in males"

**Supplementary Table 5. ANCOVA adjusted by age at sampling and APOE, using the case-control variable as an independent variable and mLRR-Y as the dependent variable.** Only individuals with available age at DNA sampling information were included in the analysis.

| | Df | Sum Sq | Mean Sq | F value | Pr(>F) |
|---|---|---|---|---|---|
| **Case control (1/0)** | 1 | 1.54 | 1.54 | 67.99 | 2.54E-16 |
| **Age at sampling** | 1 | 1.84 | 1.84 | 81.22 | 3.73E-19 |
| **APOE** | 1 | 0.09 | 0.09 | 3.86 | 4.94E-02 |
| **Residuals** | 2693 | 61.14 | 0.02 | | |
