## Supplementary Table 6 for "Mendelian randomization confirms the role of Y-chromosome loss in Alzheimer’s Disease etiopathogenesis in males"

**Supplementary Table 6. Logistic regression results for PCs asociation to AD case-control variable in the mLOY/mLRR-Y case-control analysis**. Only individuals with age at extraction 65-85 years old were included in the models.

| | log(OR) | SE | Z | P | CI2.5 | CI97.5 |
|---|---|---|---|---|---|---|
| **PC1** | -5.93 | 2.81 | -2.11 | 0.03 | -11.44 | -0.41 |
| **PC2** | -5.75 | 2.88 | -2.00 | 0.05 | -11.40 | -0.11 |
| **PC3** | 3.78 | 3.04 | 1.24 | 0.21 | -2.17 | 9.75 |
| **PC4** | -2.61 | 3.69 | -0.71 | 0.48 | -9.85 | 4.62 |
