## Supplementary Table 7 for "Mendelian randomization confirms the role of Y-chromosome loss in Alzheimer’s Disease etiopathogenesis in males"

**Supplementary Table 7. Logistic regression results for mloy-PRS, age at sampling and APOE (independent variables) vs mLOY status (dependent variable).** Only males aged 65-85 were included in the models.

| | OR | SE | z value | P | CI2.5 | CI97.5 |
|---|---|---|---|---|---|---|
| **mLOY PRS** | 1.80 | 1.07 | 9.18 | $4.22 \cdot 10^{-20}$ | 0.46 | 0.72 |
| **Age at sampling** | 1.08 | 1.01 | 6.57 | $5.07 \cdot 10^{-11}$ | 0.06 | 0.10 |
| **APOE** | 0.88 | 1.09 | -1.45 | 0.15 | -0.30 | 0.04 |
