## Supplementary Table 8 for "Mendelian randomization confirms the role of Y-chromosome loss in Alzheimer’s Disease etiopathogenesis in males"

**Supplementary Table 8. Logistic regression results for PCs asociation to AD case-control variable in the mLOY PRS case-control analysis.** Results are shown for the mixed (a), male (b) and female (c) groups. Only individuals aged 65-85 were included in the models.

a.

| MIXED (MALES + FEMALES) | logOR | SE | z.val | P | CI2.5 | CI97.5 |
|---|---|---|---|---|---|---|
| **PC1** | -0.89 | 2.32 | -0.38 | 7.02E-01 | -5.46 | 3.65 |
| **PC2** | -23.80 | 2.37 | -10.05 | 8.74E-24 | -28.46 | -19.18 |
| **PC3** | -18.71 | 2.41 | -7.77 | 7.68E-15 | -23.45 | -14.01 |
| **PC4** | 0.32 | 2.34 | 0.14 | 8.93E-01 | -4.27 | 4.90 |

b.

| MALES | logOR | SE | z.val | P | CI2.5 | CI97.5 |
|---|---|---|---|---|---|---|
| **PC1** | -1.56 | 2.38 | -0.65 | 5.12E-01 | -6.21 | 3.14 |
| **PC2** | -18.38 | 2.46 | -7.47 | 7.78E-14 | -23.23 | -13.59 |
| **PC3** | 8.62 | 2.43 | 3.55 | 3.83E-04 | 3.87 | 13.39 |
| **PC4** | -1.91 | 3.09 | -0.62 | 5.36E-01 | -7.96 | 4.14 |

c.

| FEMALES | logOR | SE | z.val | P | CI2.5 | CI97.5 |
|---|---|---|---|---|---|---|
| **PC1** | 1.71 | 2.33 | 0.74 | 4.62E-01 | -2.83 | 6.31 |
| **PC2** | -14.56 | 2.37 | -6.14 | 8.07E-10 | -19.22 | -9.93 |
| **PC3** | 12.38 | 2.37 | 5.23 | 1.72E-07 | 7.75 | 17.03 |
| **PC4** | 3.19 | 2.32 | 1.37 | 1.70E-01 | -1.36 | 7.74 |
